# First and second waves of coronavirus disease-19: A comparative study in hospitalized patients in Reus, Spain

**DOI:** 10.1101/2020.12.10.20246959

**Authors:** Simona Iftimie, Ana F. López-Azcona, Immaculada Vallverdú, Salvador Hernàndez-Flix, Gabriel de Febrer, Sandra Parra, Anna Hernández-Aguilera, Francesc Riu, Jorge Joven, Jordi Camps, Antoni Castro, REUSCOVID Study Group

## Abstract

Many countries have seen a two-wave pattern in reported cases of coronavirus disease-19 during the 2020 pandemic, with a first wave during spring followed by the current second wave in late summer and autumn. Empirical data show that the characteristics of the effects of the virus do vary between the two periods. Differences in age range and severity of the disease have been reported, although the comparative characteristics of the two waves still remain largely unknown. Those characteristics are compared in this study using data from two equal periods of 3 and a half months. The first period, between 15^th^ March and 30^th^ June, corresponding to the entire first wave, and the second, between 1^st^ July and 15^th^ October, corresponding to part of the second wave, still present at the time of writing this article. Two hundred and four patients were hospitalized during the first period, and 264 during the second period. Patients in the second wave were younger and the duration of hospitalization and case fatality rate were lower than those in the first wave. In the second wave, there were more children, and pregnant and post-partum women. The most frequent signs and symptoms in both waves were fever, dyspnea, pneumonia, and cough, and the most relevant comorbidities were cardiovascular diseases, type 2 diabetes mellitus, and chronic neurological diseases. Patients from the second wave more frequently presented renal and gastrointestinal symptoms, were more often treated with non-invasive mechanical ventilation and corticoids, and less often with invasive mechanical ventilation, conventional oxygen therapy and anticoagulants. Several differences in mortality risk factors were also observed. These results might help to understand the characteristics of the second wave and the behaviour and danger of SARS-CoV-2 in the Mediterranean area and in Western Europe. Further studies are needed to confirm our findings.

## Introduction

Coronavirus disease-19 (COVID-19), produced by the severe acute respiratory syndrome coronavirus 2 (SARS-CoV-2), has become a global pandemic, giving rise to a serious health threat globally. Several countries have seen a two-wave pattern of reported cases, with a first wave in spring and a second in late summer and autumn [1-6]. In Spain, the first wave of COVID-19 began in early March 2020, although some isolated cases had been reported in February [7]. As a consequence of the first outbreak, the Spanish Government introduced a series of strict prevention measures, including home confinement, which lasted from 13^th^ March to 4^th^ May, followed by a three-month period of progressively increasing social interaction, work and commercial activity. As of July, life in the country had returned to relative normality, except for the mandatory wearing of a face mask and maintaining a safe social distance. Unfortunately, the number of cases of patients with COVID-19 began to increase towards the end of August and a month later it once again presented numbers similar to those in April. This forced the Government to reintroduce serious restrictive measures, including local and regional lockdowns, closures of bars, restaurants, cultural and sports activities, and a general curfew after 10 pm. The second wave of COVID-19 had been predicted months earlier and had already occurred in other countries [4]. The vast majority of Western European countries are currently suffering the consequences of this second wave and are taking similar restrictive measures. However, empirical data would suggest that this second wave differs from the first in such factors as age range and severity of the disease [8]. Indeed, it has been suggested that this second wave in Europe might be linked to the appearance of a new variant of the SARS-CoV-2, termed 20A.EU1, which appears to have originated in Spain, from where it then spread to the rest of Europe through tourists who had spent their summer holidays in that area [9]. The similarities and differences between the characteristics of the two waves remain largely unknown. Population comparison is difficult because the technological and logistical capacity of the countries in detection and diagnosis of asymptomatic patients and those with mild symptoms has improved greatly in the six months since spring, and it is assumed that the incidence of infection in the early months of the pandemic was much higher than had been reported [10]. However, a more accurate comparison of the two waves is feasible through the study of the hospitalized patients for whom disease was confirmed by reverse transcription-polymerase chain reaction (RT-PCR) and severe symptoms.

This study investigated the severity and characteristics of the two waves in hospitalized patients in Reus, Spain. We evaluated age, gender, symptoms, comorbidities, mortality, supportive care, medication, and the outcome for the patient.

## Materials and Methods

### Study design

We conducted a prospective study of all hospitalized cases of SARS-CoV-2 infection in *Hospital Universitari de Sant Joan*, in Reus, Spain, admitted between 15^th^ March and 15^th^ October 2020. All patients admitted up to 30^th^ June were considered to be in the first wave and all those admitted from 1^st^ July in the second wave, which divided the study period into two equal parts of three and a half months. The only inclusion criterion was to be a hospitalized patient with an analytical diagnosis of SARS-CoV-2. We excluded those with suspected SARS-CoV-2 infection but had no laboratory confirmation and those who came to the hospital with symptoms compatible with COVID-19 but did not require hospitalization. SARS-CoV-2 infection was confirmed by RT-PCR using swab samples from the upper respiratory tract (nasopharyngeal/oropharyngeal exudate), from the lower respiratory tract (sputum/endotracheal aspirate/bronchoalveolar lavage/bronchial aspirate) or from the lower digestive tract (rectal smear). Tests were carried out with the VIASURE *SARS-CoV-2* Real Time PCR Detection Kit (CerTest Biotec, Zaragoza, Spain), or with the Procleix® method in a Panther automated extractor and amplifier (Grifols Laboratories, Barcelona, Spain). This study was approved by the *Comitè d’Ètica i Investigació en Medicaments* (Institutional Review Board) of *Hospital Universitari de Sant Joan* (Resolution CEIM 040/2018, amended on 16 April 2020).

### Calculation of sample size

Accepting an alpha risk of 0.05 and a beta risk of less than 0.2 in a bilateral contrast, it takes 137 subjects in the first wave and 105 in the second wave to detect a difference equal to or greater than 8 years in the variable age. The common standard deviation is assumed to be 22. A follow-up loss rate of 0% was estimated.

### Statistical analyses

Data is given as numbers and percentages or means and standard deviations. Statistical comparisons between two groups were made using the χ^2^ test (categorical variables) or the Student’s *t* test. Logistic regression models were fitted to investigate the combined effect of selected variables on mortality. Statistical significance was set at *p* ≤0.05. All calculations were made using the SPSS 25.0 statistical package (SPSS Inc., Chicago, IL, USA).

## Results

The raw data of this study are as Supporting Information. During the study period, 468 patients with SARS-Co-V2 infection, confirmed by RT-PCR, were admitted to the hospital. The seasonal distribution of hospital admissions is shown in Figure 1. The first wave peaked at the end of March and was followed by a progressive decrease with very few patients being admitted in May and June. The number of cases fluctuated upward from mid-July until a sharp increase in mid-October. The number of patients admitted was 204 in the first wave and 264 in the second one. Those in the second wave were significantly younger (58 ± 26 *vs*. 67 ± 18 years; p <0.001). A noteworthy feature of the second wave was the high number of children between 0 and 9 years of age (n = 21), 12 of them being babies under 1 year (Figure 2). The department to which the patients were admitted is shown in Table 1. The second wave caused a significantly higher number of admissions to Gynecology, Pediatrics and Emergency Departments and fewer to Internal Medicine and ICU. The duration of hospitalization was significantly shorter in the second wave (14 ± 19 *vs*. 22 ± 25 days; p < 0.001). A total of 49 deaths occurred during the first wave and 35 during the second wave, so the case fatality rate decreased from 24.0% to 13.2%. The patients who died were significantly older than the survivors and those who died in the second wave were older than those in the first wave (83 ± 10 *vs*. 78 ± 13 years; p = 0.042).

**Table 1.** Distribution of the hospitalized patients in the first and second waves.

| Department | First wave<br>(n = 204) | Second wave<br>(n = 264) | $p$-value |
| --- | --- | --- | --- |
| Internal Medicine | 124 (60.8) | 123 (46.6) | 0.004 |
| Intermediate Care Unit | 42 (20.6) | 47 (17.8) | 0.596 |
| Intensive Care Unit | 35 (17.1) | 19 (7.2) | 0.029 |
| Emergency Unit | 0 (0.0) | 33 (12.5) | N.A. |
| Pediatrics | 0 (0.0) | 22 (8.3) | N.A. |
| Gynecology | 0 (0.0) | 10 (3.8) | N.A. |
| Surgery | 1 (0.5) | 5 (1.9) | 0.102 |
| Oncology | 1 (0.5) | 3 (1.1) | 0.317 |
| Traumatology | 1 (0.5) | 2 (0.8) | 0.564 |

Statistical analysis was performed by the $\chi^2$ test. Results are shown as number of cases and percentages (in parenthesis). N.A.: Not applicable. The statistical test cannot be performed when one of the variables is equal to 0.

**Figure 1.**
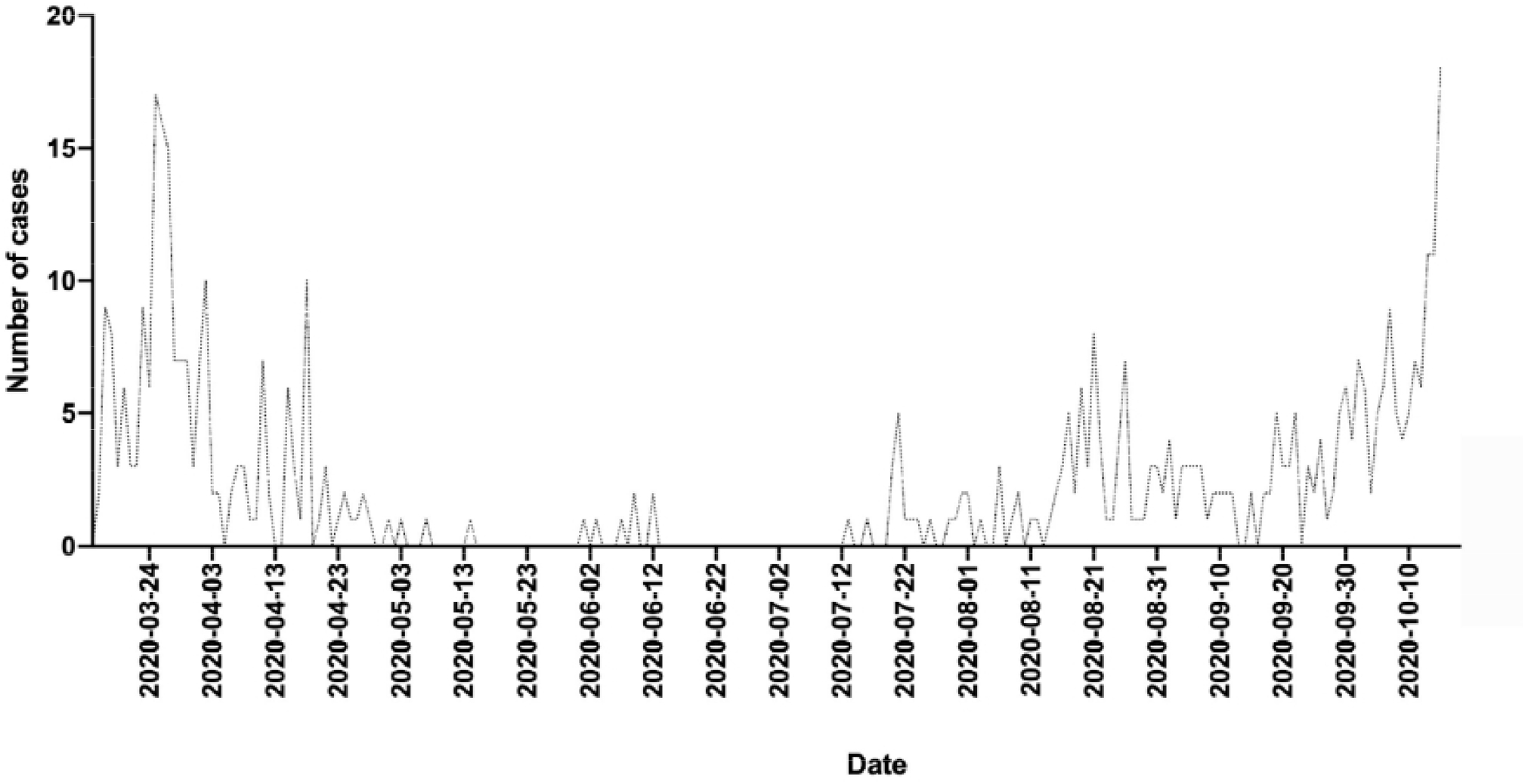
Number of patients with COVID-19 admitted per day over the entire study period.

**Figure 2.**
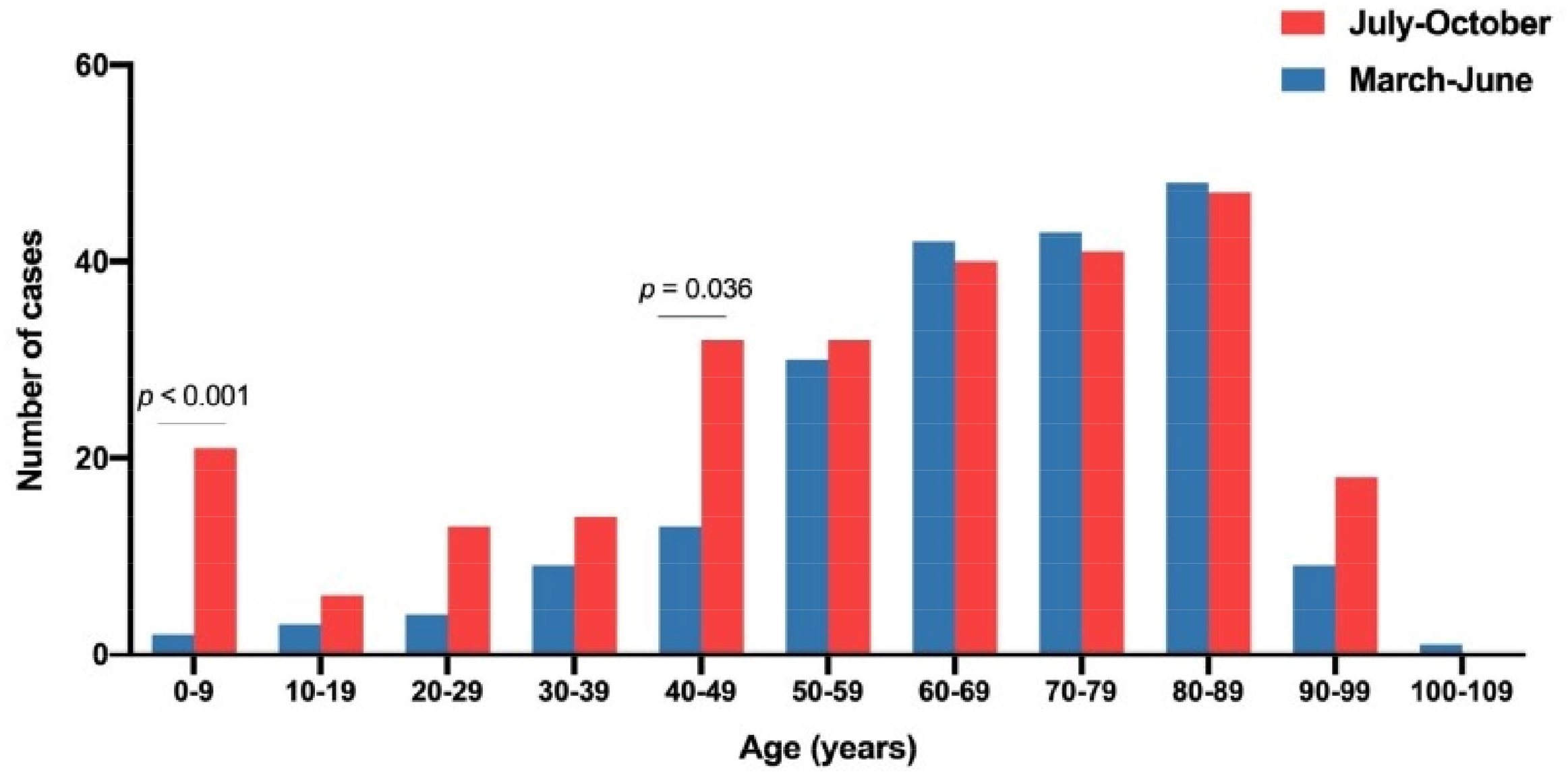
Distribution by age intervals of the patients admitted for COVID-19 during the first and second waves. The *p* values were calculated using the χ^2^ test.

The relationships between COVID-19 and the clinical and epidemiological variables are shown in Figure 3 and Table 2. The most frequent signs and symptoms in both waves were fever, dyspnea, pneumonia, and cough (Figure 3A). The most relevant comorbidities were cardiovascular diseases, type 2 diabetes mellitus, and chronic neurological diseases (Figure 3B). Patients from the second wave differed from those of the first wave in that they more frequently presented a higher frequency of vomiting, astenia, abdominal pain, rhinorrhea, or acute kidney failure, and less frequently a cough or chills. There was no significant difference in the frequency of concomitant chronic diseases. One result that we consider noteworthy is the considerably higher frequency in the second wave of pregnant women who went to the hospital to give birth and post-partum women.

**Table 2.** Clinical and epidemiological characteristics of patients with COVID-19 infection.

| Feature | First wave<br>(n = 204) | Second wave<br>(n = 264) | $p$-value |
| --- | --- | --- | --- |
| **Epidemiological characteristics** |  |  |  |
| Age | 67 ± 18 | 58 ± 26 | < 0.001 |
| Gender, male | 114 (55.9) | 144 (54.5) | 0.423 |
| Smoking habit | 10 (4.9) | 27 (13.2) | < 0.001 |
| Alcohol consumption | 10 (4.9) | 15 (7.3) | 0.421 |
| **Signs and symptoms** |  |  |  |
| Fever | 134 (65.6) | 170 (64.3) | 0.845 |
| Dyspnea | 122 (59.8) | 134 (50.7) | 0.061 |
| Pneumonia | 119 (58.3) | 140 (53.8) | 0.262 |
| Cough | 103 (50.5) | 107 (40.5) | 0.039 |
| Diarrhea | 44 (21.5) | 46 (17.4) | 0.288 |
| Chills | 42 (20.5) | 7 (2.6) | < 0.001 |
| Acute kidney failure | 22 (10.2) | 46 (17.4) | 0.048 |
| Odynophagia | 14 (6.8) | 15 (5.6) | 0.700 |
| Acute respiratory distress syndrome | 10 (4.9) | 17 (6.4) | 0.552 |
| Vomiting | 9 (4.4) | 39 (14.7) | < 0.001 |
| Other symptoms<sup>1</sup> | 12 (5.8) | 69 (26.1) | < 0.001 |
| **Comorbidities and gestational variables** |  |  |  |
| Cardiovascular disease (including hypertension) | 108 (52.9) | 144 (54.5) | 0.502 |
| Type 2 diabetes mellitus | 56 (27.4) | 64 (24.2) | 0.456 |
| Chronic neurological disease | 45 (22.0) | 52 (19.7) | 0.429 |
| Chronic kidney disease | 32 (15.6) | 34 (12.9) | 0.359 |
| Chronic lung disease | 31 (15.2) | 47 (17.8) | 0.401 |
| Cancer | 29 (14.2) | 43 (16.3) | 0.816 |
| Other infectious diseases | 6 (2.9) | 10 (3.8) | 0.464 |
| Chronic liver disease | 5 (2.4) | 17 (6.4) | 0.069 |
| Postpartum (< 6 weeks) | 2 (0.9) | 15 (5.7) | 0.024 |
| Pregnancy | 1 (0.4) | 12 (4.5) | 0.016 |

Statistical analysis was performed by the $\chi^2$ test (categorical variables) or the Student's $t$ test (quantitative variables). Results are shown as number of cases and percentages (in parenthesis) or as means ± standard deviations.
<sup>1</sup> Asthenia, rhinorrhea or abdominal pain.

**Figure 3.**
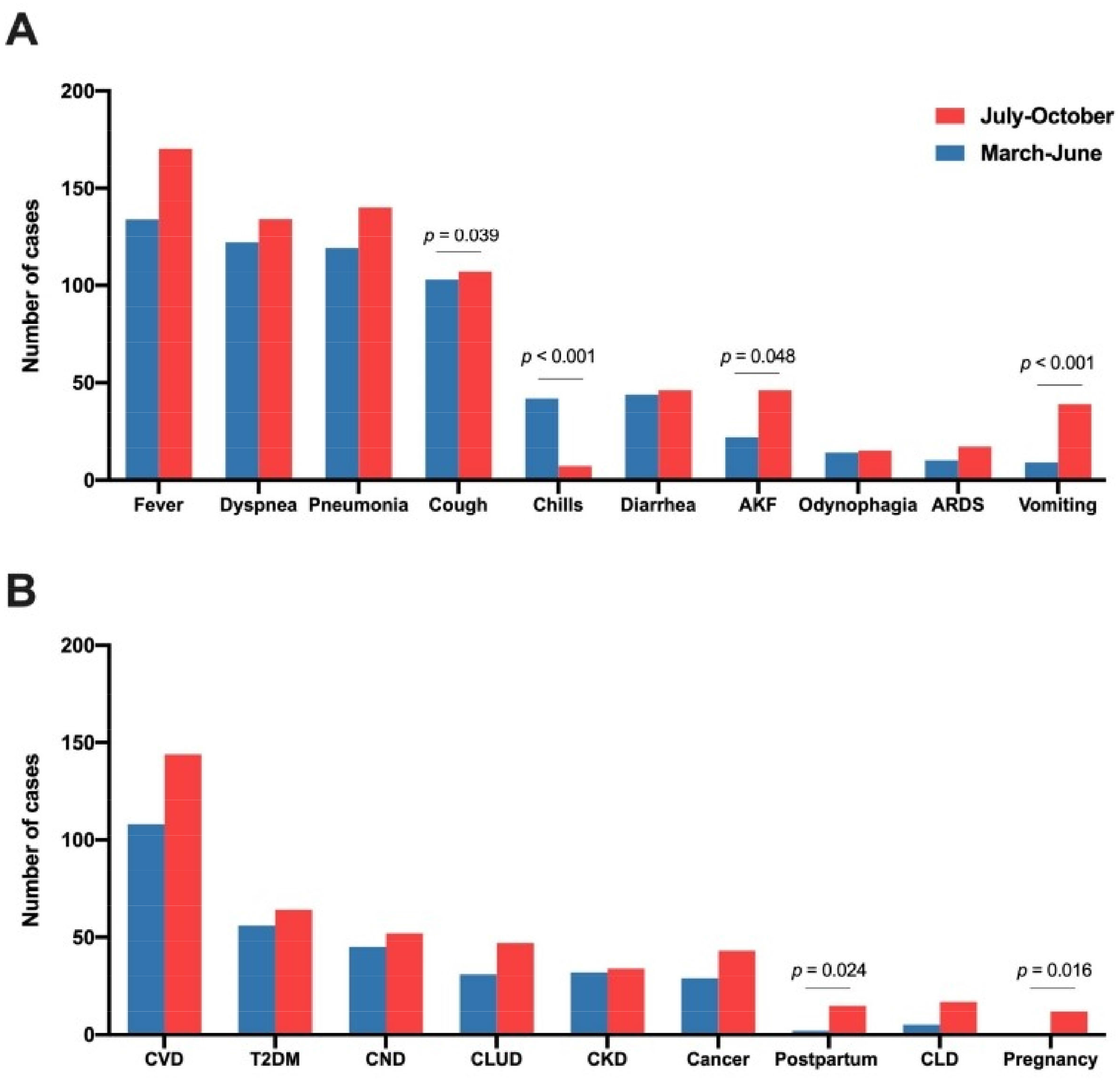
Distribution of symptoms and diseases associated with SARS-CoV-2 infection (A) and comorbidities and gestational variables (B) in patients admitted for COVID-19 during the first and second waves. The *p* values were calculated using the χ^2^ test. AKF, acute kidney failure; ARDS, acute respiratory distress syndrome; CKD, chronic kidney disease; CLD, chronic liver disease; CLUD, chronic lung disease; CND, chronic neurological disease; CVD, cardiovascular disease; T2DM, type 2 diabetes mellitus.

We also evaluated the differences in treatments between the two groups of patients. Subjects from the second wave were treated more often with non-invasive mechanical ventilation and corticoids, and less often with invasive mechanical ventilation, conventional oxygen therapy and anticoagulants (Table 3). Regarding other treatments, patients in the first wave received lopinavir, ritonavir and hydroxychloroquine, while those in the second wave received remdesivir and tocilizumab.

**Table 3.** Main treatments of patients with COVID-19 infection.

| Treatment | First wave<br>(n = 204) | Second wave<br>(n = 264) | p-value |
| --- | --- | --- | --- |
| Noninvasive mechanical ventilation | 7 (3.4) | 25 (9.5) | 0.007 |
| Invasive mechanical ventilation | 27 (13.2) | 11 (4.2) | < 0.001 |
| High-flow oxygen therapy | 18 (8.8) | 28 (10.6) | 0.315 |
| Conventional oxygen therapy | 155 (76.0) | 156 (59.1) | < 0.001 |
| Anticoagulants | 184 (90.2) | 188 (71.2) | < 0.001 |
| Corticosteroids | 86 (42.2) | 156 (59.1) | < 0.001 |

Statistical analysis was performed by the $\chi^2$ test. Results are shown as number of cases and percentages (in parenthesis).

Finally, we wanted to identify which factors were the most important determinants of death in the two groups of patients. Logistic regression analyses highlighted the importance of age, fever, dyspnea, acute respiratory distress syndrome, type 2 diabetes mellitus, and cancer in the first wave (Table 4), and of age, gender, smoking habit, acute respiratory distress syndrome, and chronic neurological diseases in the second wave (Table 5).

**Table 4.**
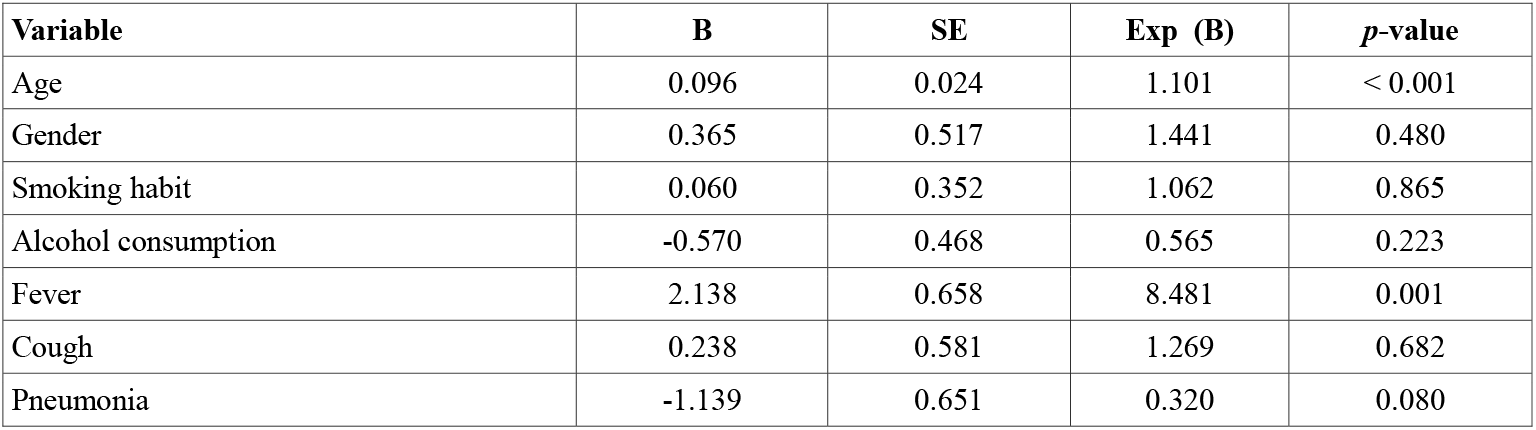
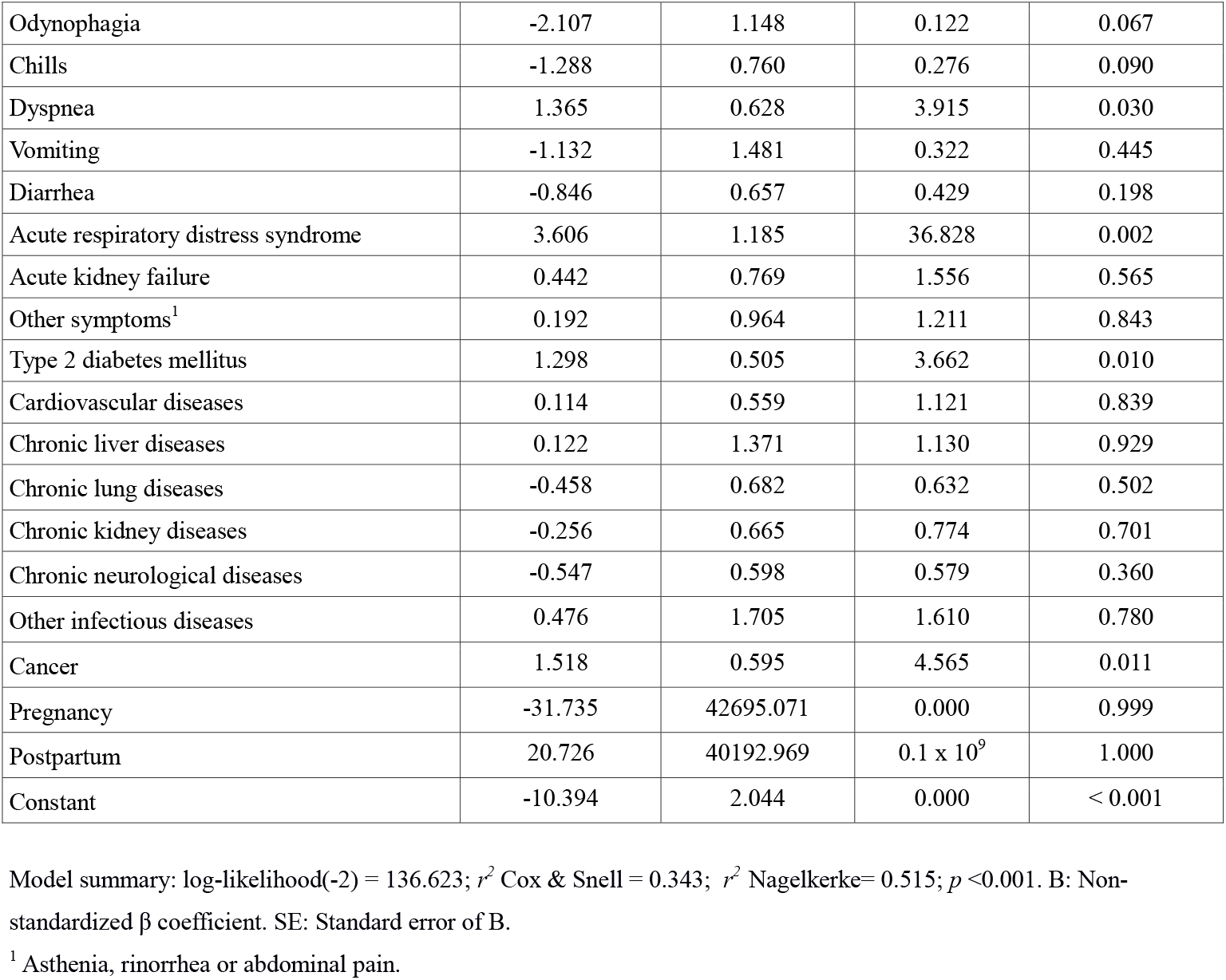
Logistic regression analysis on the relationships of comorbidities with deaths for patients from the first wave of COVID-19.

**Table 5.**
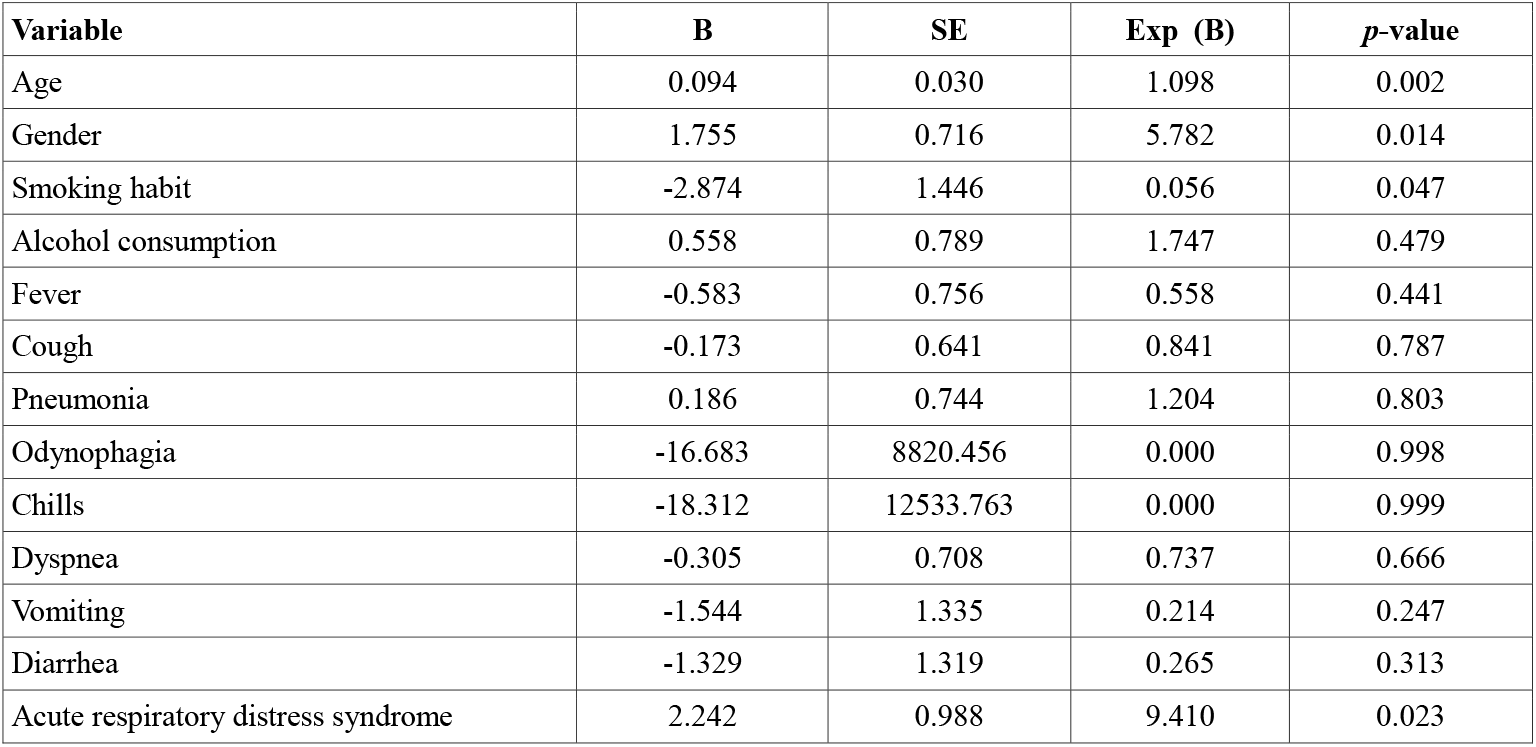
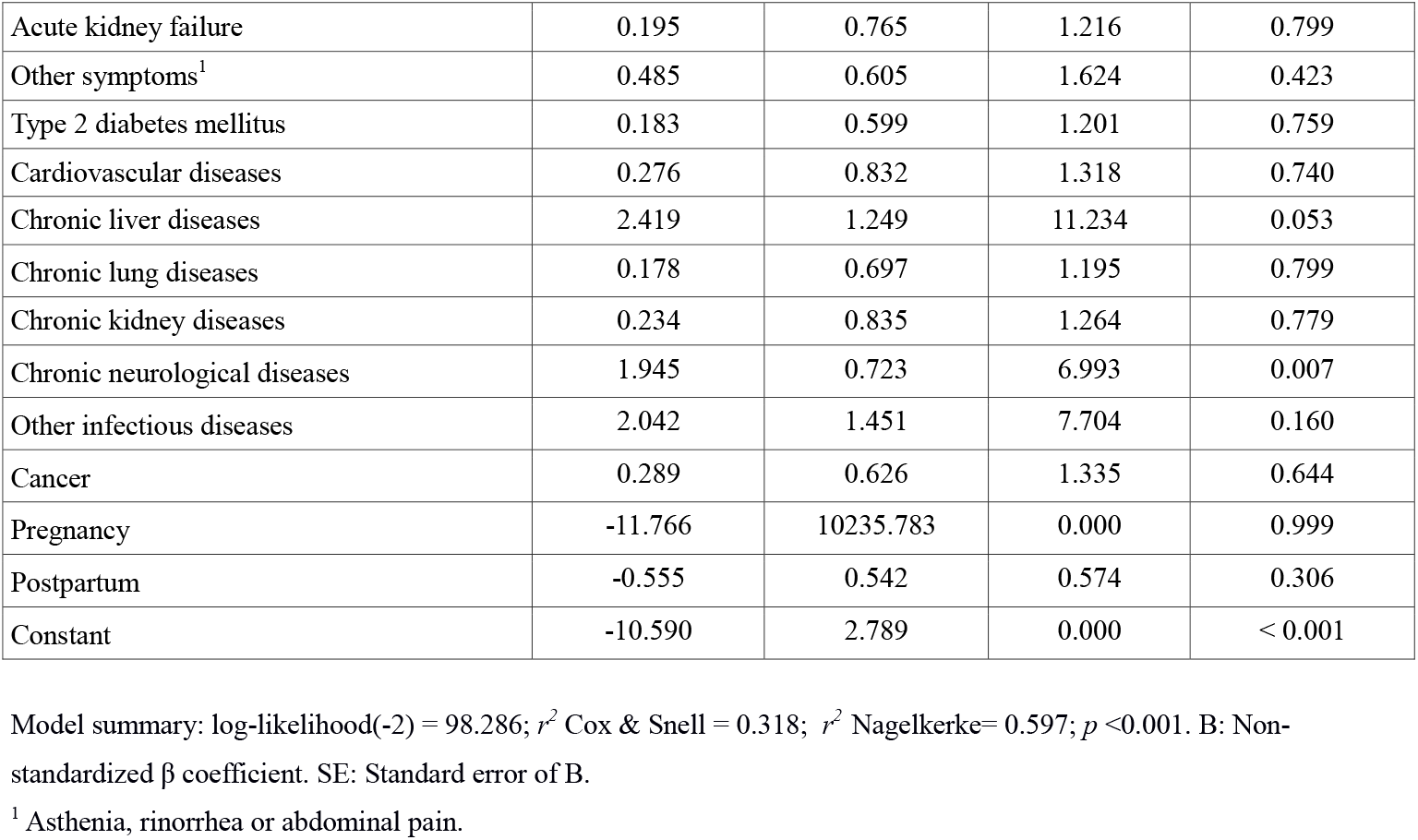
Logistic regression analysis on the relationships of comorbidities with deaths for patients from the second wave of COVID-19.

| Variable | B | SE | Exp (B) | *p*-value |
| --- | --- | --- | --- | --- |
| Age | 0.094 | 0.030 | 1.098 | 0.002 |
| Gender | 1.755 | 0.716 | 5.782 | 0.014 |
| Smoking habit | -2.874 | 1.446 | 0.056 | 0.047 |
| Alcohol consumption | 0.558 | 0.789 | 1.747 | 0.479 |
| Fever | -0.583 | 0.756 | 0.558 | 0.441 |
| Cough | -0.173 | 0.641 | 0.841 | 0.787 |
| Pneumonia | 0.186 | 0.744 | 1.204 | 0.803 |
| Odynophagia | -16.683 | 8820.456 | 0.000 | 0.998 |
| Chills | -18.312 | 12533.763 | 0.000 | 0.999 |
| Dyspnea | -0.305 | 0.708 | 0.737 | 0.666 |
| Vomiting | -1.544 | 1.335 | 0.214 | 0.247 |
| Diarrhea | -1.329 | 1.319 | 0.265 | 0.313 |
| Acute respiratory distress syndrome | 2.242 | 0.988 | 9.410 | 0.023 |
| Acute kidney failure | 0.195 | 0.765 | 1.216 | 0.799 |
| Other symptoms<sup>1</sup> | 0.485 | 0.605 | 1.624 | 0.423 |
| Type 2 diabetes mellitus | 0.183 | 0.599 | 1.201 | 0.759 |
| Cardiovascular diseases | 0.276 | 0.832 | 1.318 | 0.740 |
| Chronic liver diseases | 2.419 | 1.249 | 11.234 | 0.053 |
| Chronic lung diseases | 0.178 | 0.697 | 1.195 | 0.799 |
| Chronic kidney diseases | 0.234 | 0.835 | 1.264 | 0.779 |
| Chronic neurological diseases | 1.945 | 0.723 | 6.993 | 0.007 |
| Other infectious diseases | 2.042 | 1.451 | 7.704 | 0.160 |
| Cancer | 0.289 | 0.626 | 1.335 | 0.644 |
| Pregnancy | -11.766 | 10235.783 | 0.000 | 0.999 |
| Postpartum | -0.555 | 0.542 | 0.574 | 0.306 |
| Constant | -10.590 | 2.789 | 0.000 | < 0.001 |

Model summary: log-likelihood(-2) = 98.286; $r^2$ Cox & Snell = 0.318; $r^2$ Nagelkerke = 0.597; $p < 0.001$ . B: Non-standardized $\beta$ coefficient. SE: Standard error of B.
<sup>1</sup> Asthenia, rinorrhea or abdominal pain.

## Discussion

We have previously reported the main epidemiological and clinical characteristics and the mortality risk factors of the first wave patients during a month and a half between March and April [11]. In the present investigation we extended the study to mid-October to cover two equal periods of three and a half months. More patients were admitted during the second wave, they were younger and there were fewer deaths, in agreement with results reported by previous research in several countries [2,3,12]. The reasons for the clear differences between the two periods are not yet known although it has been suggested that a new variant of SARS-CoV-2 emerged in early summer 2020 in Spain [9], a variant that was linked to outbreaks among young agricultural workers in the north-east of the country. Transmission to the general population in that area was then replicated across the country. Furthermore, poor compliance with social distancing guidelines by young people might have facilitated contagion in young, healthy adults and children [2,13]. The decrease in the age of the patients then resulted in a decrease in the case fatality rate in that those patients who died were on average 5 years older than the victims of the first wave. Moreover, fewer patients required respiratory assistance via invasive mechanical ventilation methods. This improvement in the results of admitted patients might be linked to the fact that the health system in our country, as in many others, has since become better prepared. We have more experience and better treatment regimens, and we carry out more diagnostic tests, allowing serious cases to be detected early and to receive more effective treatments. In this regard, during the second period, patients were treated more frequently with dexamethasone, as suggested by the RECOVERY study [14], and hydroxychloroquine and loponavir-ritonavir were substituted by remdesivir and tocilizumab, which several studies have reported to be more effective than in preventing death and shortening the duration of hospital stays [15-17]. Another factor that might have contributed to the decrease in the case fatality rate is the improvement in environmental conditions. For example, warm weather and improved air quality following the city lockdown have been reported to correlate negatively with SARS-CoV-2 transmissibility [18-20].

A new and remarkable characteristic of the incidence of COVID-19 in this second wave in our population is the higher incidence in babies, children and pregnant women who went to the hospital to give birth or in post-partum women. The vast majority of these patients did not present serious symptoms and so did not require hospitalization for more than 4 days. There were no deaths among children up to 9 years of age, pregnant or post-partum women. The predominant symptom presented by the children was fever (19 out of 21 cases, 90.5%), while pregnant and post-partum women (13 and 17 cases, respectively) were asymptomatic and promptly discharged. These results highlight the role of family contact in the transmission of the virus and agree with previous reports that have indicated the generally low severity of the disease in these patients [21-24].

The predominant symptoms of infection (fever, dyspnea, pneumonia cough) were similar in both waves, although the patients in the second wave presented renal (acute kidney failure) and gastrointestinal symptoms (vomiting, abdominal pain) more frequently. Indeed, the Spanish Ministry of Health has already highlighted, in a document updated on 2^nd^ October, the higher incidence of the latter in the second wave [25]. The present study did not find any differences between the frequency of concomitant diseases in the two waves, similar findings to those of our preliminary study [9]. In this respect, we differ from a previous study conducted in Japan that has reported a lower incidence of cardiovascular and cerebrovascular diseases [3].

Lastly, regarding the risk factors associated with mortality, we also found differences between the first and second waves. Multiple regression analysis showed that older age and the presence of fever, dyspnea, acute respiratory distress syndrome, diabetes, and cancer were independently associated with higher mortality in the first wave, while age, gender, and the presence of acute respiratory distress syndrome and chronic neurological diseases were associated with mortality in the second. This might be a reflection of a better management of cancer or diabetes patients. On the other hand, the association of neurological diseases with mortality might be due to the higher mean age of those who died in this second wave.

## Conclusion

The results of the present study show that hospitalized patients in the second wave were younger, required fewer days of hospitalization, had lower mortality rates and treatments were more effective and less intensive. Although the majority of symptoms were similar in both periods, the higher incidence of gastrointestinal symptoms in the second wave stands out as a difference. Comorbidities were similar, but there were differences between those associated with mortality, highlighting the importance of chronic neurological diseases in this second wave. An important difference was the high incidence of babies, children and pregnant and post-partum women admitted but, in general, these cases were not serious and were resolved promptly and successfully. These results might help to understand the characteristics of this second wave and the behaviour and danger of SARS-CoV-2 in the Mediterranean area and in Western Europe generally. Further studies are needed to confirm our findings.

## Data Availability

All relevant data are within the manuscript and its Supporting Information files.

## Acknowledgments

The authors are indebted to all the staff of the *Hospital Universitari de Sant Joan*, doctors, nurses, assistants, cleaning and security personnel, and all the volunteer students, who with their enormous effort are managing to overcome this dramatic situation. Editorial assistance was provided by Phil Hoddy at the Service of Linguistic Resources of the *Universitat Rovira i Virgili*.

## CRediT authorship contribution statement

**Simona Iftimie:** Conceptualization, Data curation, Formal analysis, Investigation, Methodology, Project administration, Resources, Supervision, Validation, Funding acquisition, Writing-original draft, Writing-review & editing. **Ana F. López-Azcona**: Data curation, Investigation, Methodology, Writing-review & editing. **Immaculada Vallverdú:** Investigation, Resources. **Salvador Hernández-Flix:** Investigation, Resources. **Gabriel de Febrer:** Investigation, Resources. **Sandra Parra:** Investigation, Resources. **Anna Hernández-Aguilera:** Data curation, Investigation, Methodology, Writing-original draft, Writing-review & editing. **Francesc Riu:** Investigation, Resources. **Jorge Joven:** Investigation, Resources. **Jordi Camps:** Conceptualization, Data curation, Formal analysis, Investigation, Methodology, Project administration, Resources, Supervision, Validation, Funding acquisition, Writing-original draft, Writing-review & editing, Supervision. **Antoni Castro:** Investigation, Resources, Funding acquisition, Writing-review & editing.

## Data availability

All relevant data are within the manuscript and its Supporting Information files.

